# Impact of COVID-19 restrictions on pre-school children’s eating, activity and sleep behaviours: a qualitative study

**DOI:** 10.1101/2020.12.01.20241612

**Authors:** JL Clarke, R Kipping, S Chambers, K Willis, H Taylor, R Brophy, K Hannam, SA Simpson, R Langford

## Abstract

**Background:** In spring 2020, the COVID-19 lockdown placed unprecedented restrictions on the behaviour and movements of the UK population. Citizens were ordered to ‘stay at home’, only allowed to leave their houses to buy essential supplies, attend medical appointments or exercise once a day. This qualitative study explored how ‘lockdown’ and its subsequent easing changed young children’s everyday activities, eating and sleep habits to gain insight into the impact for health and wellbeing.

**Methods:** In summer 2020 we interviewed 20 parents of children due to start school in September 2020 (aged 3-5 years) by phone or video call to explore their experiences of lockdown and its easing. We recruited participants through nurseries and local Facebook community groups in the South West and West Midlands of England. Half the sample were from Black, Asian or Minority Ethnic backgrounds and half lived in the most deprived quintile. We analysed interviews using thematic analysis.

**Results:** Children’s activity, screen time, eating, and sleep routines had some level of disruption. Parents reported children ate more snacks during lockdown, but also spent more time preparing meals and eating as a family. Most parents reported a reduction in their children’s physical activity and an increase in screen time, which some linked to difficulties in getting their child to sleep. Parents sometimes expressed guilt about changes in activity, screen time and snacking over lockdown. Most felt these changes would be temporary with no lasting impact, though others worried about re-establishing healthy routines.

**Conclusions:** The spring COVID-19 lockdown negatively impacted on pre-school children’s eating, activity and sleep routines. While some positive changes were reported, there were wide-spread reports of lack of routines, habits and boundaries which, at least in the short-term, were likely to have been detrimental for child health and development. Guidance and support for families during times of COVID-19 restrictions could be valuable to help them maintain healthy activity, eating, screen-time and sleeping routines to protect child health and ensure unhealthy habits are not adopted.

## Background

The emergence of COVID-19, and subsequent efforts to restrict its spread, has led to unprecedented economic and social disruption across the globe [1]. Most affected countries have imposed restrictions on citizens’ behaviours and movements to limit the spread of the virus, including social distancing and imposing local or national ‘lockdowns’.

The UK Government imposed the spring ‘lockdown’ on 23^rd^ March 2020. UK citizens were required to ‘stay at home’ only leaving to buy essential supplies, attend medical appointments or to exercise. Schools and nurseries were closed (except to vulnerable children or children of ‘keyworkers’), as were leisure facilities (including playgrounds), pubs, restaurants, theatres and non-essential shops. Non-essential workplaces were closed, and people could not meet anyone beyond their immediate household. Unlike more stringent lockdowns (e.g. Spain and Italy), the UK population were permitted to leave the house once a day to exercise, recognising the physical and mental health benefits this provided [2]. Restrictions lasted over 2 months until their gradual easing in late May 2020. The UK government asked pre-schools to reopen to all children on 1 June 2020, alongside schools who re-opened to certain year groups at this point; however, occupancy remained low with attendance half that recorded for the same period the previous year (37% vs.77%) [3].

Despite apparently being least affected by the virus [4]children have experienced enormous disruption to their everyday lives. These changes may have had a significant impact on children’s health behaviours. Children under five should engage in 180 minutes of physical activity a day [5]. Children aged 3-4 should have between 11 hours and 12 hours 45 minutes sleep in a 24-hour period [6]. Yet the ‘stay at home’ order reduced children’s opportunities for physical activity, while increasing the likelihood of sedentary behaviours [7] and had the potential to disrupt sleep patterns [8]. The spring lockdown also brought significant changes to people’s food shopping and eating habits. Consumers moved to less frequent shopping trips, but often spent more on food than pre-lockdown, potentially changing the home food environment [9].

Several rapid surveys have sought to capture the impact of COVID-19 on children and young people’s health and well-being. Most have focused on emotional and mental health and have often targeted older children and young people, nationally and internationally [10]. Less research attention has focused on younger (pre-school) children or explored the impact on health behaviours such as physical activity, sedentary behaviour and dietary intake. An exception is the Co-SPYCE study [11] which surveyed caregivers of 2-4 year olds. It found 26% of children had three or more hours of screen time a day, and only 22% were meeting the target of 180 minutes physical activity a day. Public Health Scotland developed the COVID-19 Early Years Resilience and Impact Survey (CEYRIS) [12] to understand the impact on 2-7 year olds in Scotland. From 11,228 participants, eating behaviour was identified as worse by 32% of parents. There were mixed responses in the CEYRIS about physical activity, with 24% of parents reporting children doing more during lockdown and almost half (47%) reported less.

This study explored the impact of lockdown, and its easing, on pre-school (3-5 years) children through qualitative interviews with parents focusing on: physical activity, sedentary behaviour, food intake and sleep. Establishing healthy behaviours in the early years is important in maintaining a healthy weight throughout childhood and beyond [13]. Nearly one in four UK children starting primary school are overweight or obese, with rates increasing with deprivation[14]. Many children saw time in childcare reduced or stop altogether [15]. Pre-school aged children require greater parental supervision than older children which may mean managing children’s activities and behaviours in lockdown was more challenging for these families, particularly if parents were working from home or there were additional stressors for the family [16].

## Methods

### Population, sampling and recruitment

Criteria for inclusion were: (1) parent/carer of a child aged 3-5 years in their final year at pre-school; child usually attends pre-school at least once per week; (3) child was due to start school in September. Non-English speakers were excluded for pragmatic reasons.

Parents were recruited via two methods. First, we asked nurseries participating in another research study [17] to disseminate study information to parents via email or social media. This included nurseries from Swindon and Somerset (South West England) and Sandwell (West Midlands). Second, we posted study information on local Facebook groups within these areas and Birmingham (West Midlands) to increase recruitment of Black, Asian and Minority Ethnic (BAME) participants. Interested parents (n=85) completed an online form to check eligibility and collect postcode (for calculation of Index of Multiple Deprivation (IMD) score) and ethnicity data for sampling purposes. To provide socioeconomic and ethnic diversity in the sample, sampling was weighted towards parents in the most deprived areas and those from BAME populations. Twelve parents were recruited via nurseries and eight via Facebook.

Sampled parents were emailed the participant information sheet (PIS) and consent form. A convenient time to conduct the interview was arranged with participants at least 24 hours after sending study documents. All sampled parents agreed to participate, bar one who we were subsequently t unable to contact.

### Data collection

Semi-structured interviews were conducted by experienced qualitative researchers (JC, KW or SC) in July/August 2020. We conducted telephone (n=18) or video (n=2) interviews, at the participant’s preference. Before interview, the researcher explained the purpose of the research, answered any questions and gained verbal consent. A topic guide (Table 1) was used flexibly to guide discussions with parents encouraged to talk openly about their experiences. Interviews were audio-recorded, transcribed verbatim and anonymised. Following each interview, notes were made of key points raised. A £30 shopping voucher was sent to participants following interview.

**Table 1.**
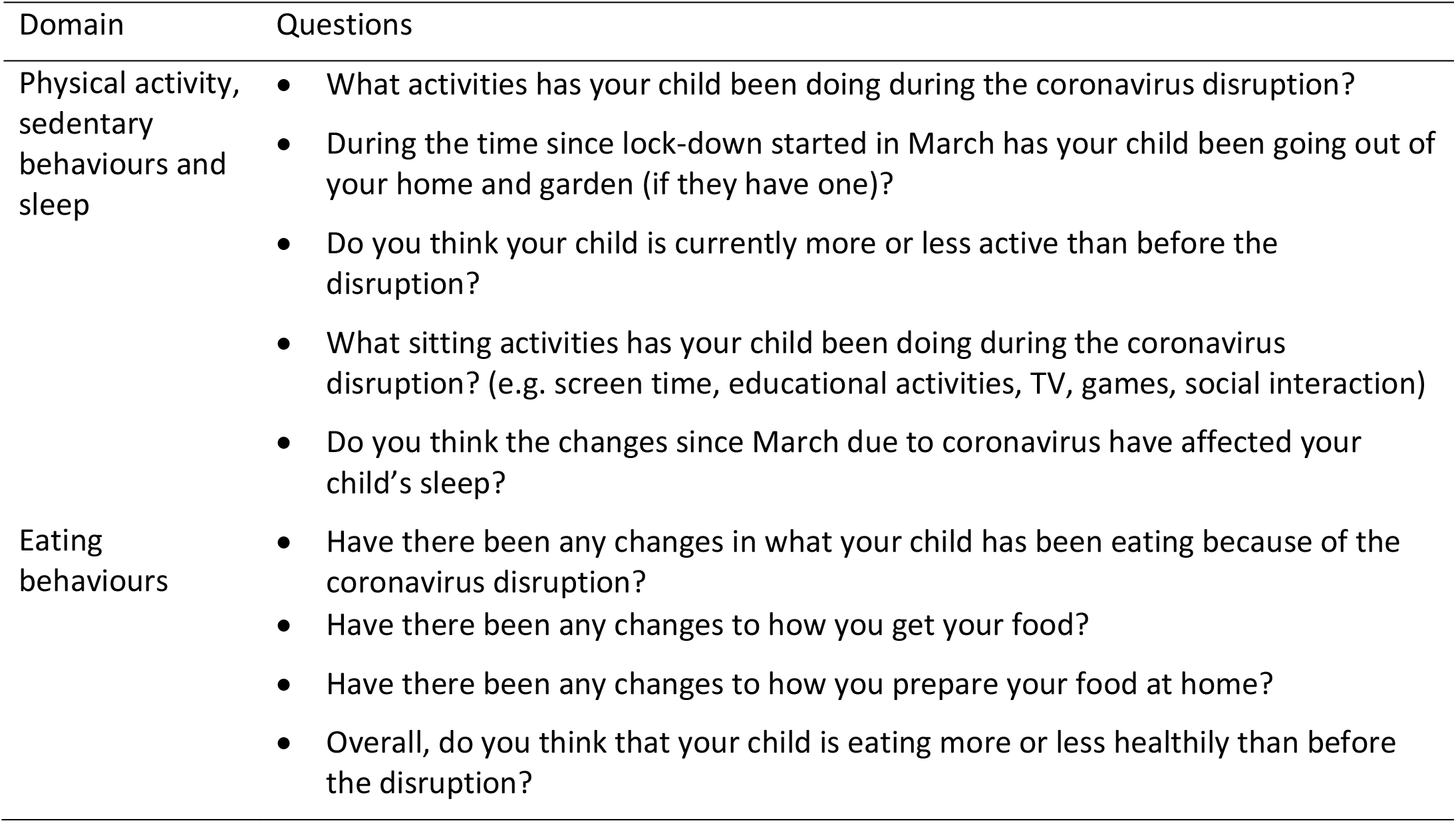
Topic guide used by interviewers to guide discussions during semi-structured interviews with parents of pre-school children.

| Domain | Questions |
| --- | --- |
| Physical activity, sedentary behaviours and sleep | • What activities has your child been doing during the coronavirus disruption?<br>• During the time since lock-down started in March has your child been going out of your home and garden (if they have one)?<br>• Do you think your child is currently more or less active than before the disruption?<br>• What sitting activities has your child been doing during the coronavirus disruption? (e.g. screen time, educational activities, TV, games, social interaction)<br>• Do you think the changes since March due to coronavirus have affected your child's sleep? |
| Eating behaviours | • Have there been any changes in what your child has been eating because of the coronavirus disruption?<br>• Have there been any changes to how you get your food?<br>• Have there been any changes to how you prepare your food at home?<br>• Overall, do you think that your child is eating more or less healthily than before the disruption? |

### Data analysis

Transcripts were checked for accuracy before analysis. Data were analysed thematically [18] using NVivo version 12 software (QSR International) to aid data management and analysis. Transcripts were read by JC/RB/HT to gain familiarity with the data. Three transcripts were independently coded by these researchers to generate an initial list of codes. Codes were both deductive (generated from our topic guide and research questions) and inductive (generated from interview data). Differences in coding were resolved through discussion to produce an agreed coding framework. Subsequent transcripts were single-coded using this coding framework with further discussion to clarify or expand the framework as needed.

### Ethical considerations

Ethical approval was granted by University of Bristol Faculty of Health Sciences Research Ethics Committee (ref: 106002).

## Results

### Participants

Sixteen mothers and four fathers participated in the study (Table 2). The average age of parents was 34 years (range 21-45 years). Half the sample described their ethnicity as White British. Thirteen participants were educated to degree level. Half resided in the most deprived IMD quintile. Five participants were single parents. Thirteen participants reported at least one parent in their household was not working during the lockdown period; a further three reported that no household member was currently in employment. Two participants did not have access to a garden.

**Table 2.** Demographic characteristics of the parents of pre-school children who participated in a semi-structured interview (n=20)

| Characteristic Categories |  | N(%) |
| --- | --- | --- |
| Gender | *Female* | 16 (80) |
|  | *Male* | 4 (20) |
| Age Group | *21-25* | 1 (5) |
|  | *26-30* | 7 (35) |
|  | *31-35* | 5 (25) |
|  | *36-40* | 2 (10) |
|  | *41-45* | 5 (25) |
| Ethnicity | *White* | 12 (60) |
|  | *Asian or Asian British* | 4 (20) |
|  | *Black or Black British* | 1 (5) |
|  | *Mixed* | 3 (15) |
| IMD Quintile<br>(1 = most deprived) | *1* | 10 (50) |
|  | *2* | 2 (10) |
|  | *3* | 3 (15) |
|  | *4* | 3 (15) |
|  | *5* | 2 (10) |
| Anyone in the household<br>in paid employment | *Yes* | 17 (85) |
|  | *No* | 3 (15) |
| Educated to degree level | *Yes* | 13 (65) |
|  | *No* | 7 (35) |
| Single parent | *Yes* | 5 (25) |
|  | *No* | 15 (75) |

### Food and eating

#### Increased snacking

Most parents reported their child’s snacking had increased over lockdown, something they often linked to the stay-at-home restrictions. Children were bored and consequently snacked more as they were *“cooped up”* at home. As Mother_1 explained, “*When there’s nothing to occupy them, they’ve got to do something”*. Others felt loss of routine had disrupted rules around snacking: “*if he’s at pre-school he can’t snack. He can only eat at the specific snack time and the specific lunchtime. Here, he just thinks it’s on tap*.*”* (Mother_2). Being together all day made dealing with demands for snacks challenging, with some parents allowing greater access to less healthy items than normal. Mother_3 explained, *“It’s all we hear. She’s constantly hungry… She’s definitely eating more bags of crisps than she normally would”*.

In response, some parents created make-shift rules around food, balancing child demands with wanting to provide ‘healthy’ snacks. Mother_4’s children were allowed crisps because they went *“on and on at me*” but then were told “*to move on to the fruit bowl*.*”* Another parent made healthy snacks more accessible: “*I make sure there are bits and bobs cut up in the fridge. Because I know if she’s going to help herself to something, it’s the stuff on the bottom she can reach”* (Mother_5). However, some parents reported giving additional ‘treats’ to their children to compensate for the pandemic’s impact: *“We were feeling bad they’d missed out on so much, we were buying treats and things for them to make them feel happy*.” (Mother_6)

### Food preparation and mealtimes

Some parents reported an increase in problematic behaviours at mealtimes, such as children not wanting to sit down for meals or taking a long time to eat. Often this was linked to their child’s increased snacking. However, several reported positive effects of lockdown on mealtime routines and behaviours. With parents working from home, families could eat together, which both parents and children appeared to appreciate. Similarly, parents enjoyed having extra time to cook and involve their child in food preparation. This helped occupy their child occupied and taught them new skills: “*When we couldn’t go to McDonald’s we made homemade chicken nuggets… that’s been posi*tive, *they’ve enjoyed cooking”* (Mother_6).

Lockdown had other impacts on families’ food behaviours. With restaurants shut, people no longer ate out. Some participants also reported consuming less take-aways. For one, this was related to concerns about the virus – “*I didn’t touch any takeaways. I did not feel comfortable to do so*.*” (*Mother_4). In contrast, one parent reported eating more take-aways because “w*e all need something to look forward to”* (Mother_7).

### Food costs

Several participants reported spending more on food during lockdown. Some related this to the whole family now being at home, as well as increased snacking, while others felt prices increased. Lockdown increased family food bills in other ways too. One family no longer had access to free meals at their child’s nursery: “*It’s a lot more expensive now obviously, because she was having meals* [at pre-school]… *We haven’t had any kind of help from school with regard to her dinner”* (Mother_8). Another participant who was shielding had moved to on-line deliveries, a service unavailable at their normal cheaper supermarkets: “*We’d normally just go to Aldi or Lidl and be able to buy it, whereas now we’ve been trying to do the online stuff, it’s just added to our bill*” (Mother_9). However, an unexpected benefit of this was healthier snacking: “*it’s a money thing, we’ve not been able to buy all the junk food …The stuff he can see is the fruit bowl, so that’s probably improved a bit*” (Mother_9).

### Physical activity

Most parents felt their child was less active during lockdown. Some perceived this reduction to be substantial - “*we* [were] *nowhere near as active as we used to be”* (Mother_4) *–* while a minority felt it had little impact – *“She’s always on the go anyway”* (Mother_5). Several parents related this reduction to the closing of childcare facilities which removed usual opportunities for activity: *“I put it down to routine and being less active… At nursery, you’re on the go from the time you get there to the time you go home”* (Mother_10). Opportunities for active transport to and from childcare were also lost: “*[She’s] definitely less active because we walk to [pre-]school and that was 10 minutes there and 10 minutes back… She’s not having that exercise*” (Mother_11). In addition, normal activities (like swimming, soft play) were closed and opportunities to socialise with friends or grandparents were restricted.

Parents noted several factors that helped children stay active. Having a sibling was important, providing someone to play with. Equally, access to play equipment helped facilitate activity: *“They’ve got a trampoline, a bike each, a scooter each*… *a climbing frame”* (Mother_4). Access to outside space was also important. Only two parents had no access to outdoor space, but both described this as challenging s during lockdown. As Mother_8 explained, “*While we were in the main period of lockdown, we couldn’t leave and there was nothing really that you could do. And living in such a block of flats, you can’t be too noisy because you’ve got neighbours everywhere*.”

In contrast, Mother_1 noted how having a safe space to cycle outside her house helped keep her son active: “*My front* [garden] *is really nice for children to play and ride bikes… And biking is one exercise* [my son] *really loves doing*.” The good weather experienced during much of lockdown was also seen as an important facilitator of activity: “*We had the pool out and [he] was using it quite a lot … We were pretty much burning energy off most of the time really, especially with the nice weather we had*” (Mother_12). Some parents discussed how they made use of the local environment during lockdown. *“We’ve done a lot of exploring around the local area. We don’t like to go too far, just in case, but we’ve found some places we didn’t know were there”* (Mother_6).Parents often put effort into keeping their child physically active: *“I’ve tried really hard to make sure he’s been as active as possible”* (Mother_7). This was easier for families where one parent was not working: “*He has to have a parent with him. ‘Look at me! Look at my star jumps!’… That was the difficult bit, because there is only so much of that you can do, especially when you’ve got work*.” (Father_1)

### Sedentary behaviours

Parents reported their child had engaged in a range of sedentary activities during lockdown and its easing, including reading, drawing and crafting. However, screen time was mentioned most frequently with almost all parents reporting substantial increases. For many families, screen time filled the void left by being unable to go out or socialise. Mother_13 explained they watched more television because “*we were stuck in the house… we weren’t having that release of getting out”*, while Mother_1 commented “[My son] *can’t go to friends, he can’t meet family… so the screen had become his friend”*.

However, many felt screen time had been useful during this difficult period, with one parent referring to it as a *“lifesaver”* (Father_1). For those who were trying to work from home, it was often the only way they could manage: “*If I’ve got a two-hour Zoom meeting and I don’t want him bursting in every five seconds I’d give him his iPad and he’ll watch a film for two hours… it’s been the only way to make sure he’s safe*” (Mother_6). Even for those not working, managing everyday chores without their usual childcare was a challenge: *“You’ve still got housework to do, washing to do, meals to prepare… You can’t always be there 24/7 to keep her occupied”* (Mother_3).

Screen time was also used to provide much needed respite from the intensive parenting effort lockdown enforced. Father_2 described screen time providing “*a break”* while Mother_11 used it to avoid constant sibling squabbles: *“They’re just following you around … fighting and hitting each other… just sit down quietly and watch a movie now*.*”*

However, parents’ language suggested many felt uncomfortable about this increased screen time. Mother_6 admitted *“It’s bad, I don’t even like saying it out loud, but [screen time] was almost like it was a pacifier, really*”, while Mother_3 admitted “*It’s going to sound horrible, but it’s just sometimes easier”*.

Parents often distinguished between ‘good’ and ‘bad’ screen time. Educational or interactive screen time was considered better than passive television watching. Mother_11 for example would ask herself “*Is it educational or is it just rubbish they’re watching?”*, while Mother_3 explained “*I don’t just want her to mindlessly sit there watching a screen. At least if she was playing [educational] games, it’s still making her think*.”

Concerns over screen time were rarely linked to lack of movement or activity. Rather Father_3 referred to *“the effect of the brain becoming lazy”* which he admitted was *“something that is worrying me*.*”* Mother_5 suggested too much screen time interfered with normal childhood and fostered an inability to occupy oneself: “*They’re not being children. I suppose you think about your own childhood, don’t you? And all the bits and bobs I used to do as a kid. When I’ve said no screen time, they just all wander around like aimless sheep*.”

Some parents created new household rules to manage this increased screen time. This included restrictions on the amount, times of day or content of screen time. Mother_14 explained, “*We have had to impose rules… we’ve just had to structure it a bit more, because she would gladly sit in front of the telly or iPad all day*.” Other families took a more reactive approach: “*We’re not a routine sort of family anyway, so if they’ve had an hour or so, then I’ll be like, ‘Right, that’s it now, everybody out*.*’”* (Mother_5)

### Sleep

Almost half of parents reported negative changes to their child’s sleep. Many reported difficulties in getting their child to sleep, with some staying up very late. Parents often related this either to lack of activity – “*I should just imagine it’s because she’s not been as active and been out*” (Mother_8) – or increased screen time – “*a direct correlation of being a bit more bored, spending more time on a screen”* (Mother_3). Loss of routine associated with lockdown was again a source of difficulty for some families. As Mother_4 commented: “*Whereas before it was a very strict* [bedtime] *routine* … *now it is kind of as and when”*. A minority of parents wondered if anxiety played a part in their child’s sleep difficulties: “*I’m just wondering whether [*his frequent waking*] was a part of maybe anxiety or worried about everything that’s going on or a phase that he went through*.” (Mother_12)

### Longer term impacts

Generally, parents thought children’s increase in snacking during lockdown was “*just a temporary thing”* (Mother_2) and would not last once children were back at nursery or school. As Mother_5 acknowledged: “*when she goes to school she’s not going to have that opportunity to (a) steal food or (b) she won’t be home to be eating masses during the day”*. However, one parent whose child had already returned to nursery acknowledged getting back into a routine was difficult: *“Trying to get him to eat properly once we started to go back into a routine was really hard* (Mother_10).

Several parents wished to sustain positive changes established during lockdown, such as home-cooked meals and involving children in food preparation. However, as one mother noted, these changes required time and would need a *“conscious effort to keep doing”* (Mother_14). Many talked positively about returning to previous activities (e.g. swimming, gymnastics) and were pleased to see these opening again. However, several admitted feeling *“wary”* (Father_4) about how safe these were, especially those involving close contact (such as soft play).

Some parents expressed concerns about the impact of lockdown on their child’s skills and confidence. Mother_14 felt her daughter would have lost her swimming skills over lockdown and noted a loss of confidence in other areas too: “*She’s a lot more cautious on her bike because for a while they just didn’t leave the house… she’s had a step backwards with that*.” Another parent was concerned about lost stamina and wondered if her daughter would manage the mile-long walk to school: “*I am actually a little bit worried about how she’s going to fare… she’s not been used to doing that walk*” (Mother_5).

Most parents were not overly concerned about increased screen time, assuming it would naturally decrease with a return to routine: “*I should imagine when she goes back that will just resume back to normal” (*Mother_8). However, others wondered how an increased reliance on screens during lockdown might change their child’s behaviours going forwards. Mother_6 described her son pre-lockdown as a *“pocket rocket*” who never sat still. Now, however, “*rather than going outside and running around the garden, as soon as he wakes up [he asks] ‘Can I have the iPad?’…He would quite happily sit all morning without moving*.” Similarly, Mother_3 explained her daughter had *“definitely got to a point where she would rather have something on the telly than go out”*. Allowing her daughter to watch television in her mother’s bedroom during lockdown, Mother_3 acknowledged the difficulty in re-establishing pre-lockdown boundaries: “*She’s asking Santa for a television for her bedroom, which I’m hoping she forgets*.”

## Discussion

During COVID-19 lockdown parents reported increased snacking, screen time and sleep issues, and decreased physical activity. There were also some positive family behaviour changes, such as more eating together and use of the local environment for physical activity. Although some parents expressed concerns about the future impact of negative behaviour changes, most believed changes were a temporary consequence of a lack of routine.

This study provides in-depth reports of family life with a pre-school child during the pandemic, and thus complements survey findings of parents of 2-4 year olds from the Co-SPYCE study [11] which revealed children’s screen time as a key parental stressor during lockdown. Our finding of increased children’s screen time and the associated parental guilt resonates with Co-SPYCE findings. Parental use of children’s screen time to ‘get things done’ was a key finding of a recent qualitative systematic review [19]. In our study, with the absence of childcare, and corresponding increase in parental juggling of work and household chores, it is clear how this ‘need’ for downtime increased for many parents.

Most parents felt their child’s physical activity had decreased over lockdown echoing findings from other studies. Only 22% of parents in the Co-SPYCE study reported their 2-4 years olds were meeting the UK recommendation of at least three hours physical activity per day [20] and almost half (47%) of Scottish parents surveyed in the CEYRIS reported their children (2-7yrs) did less physical activity during lockdown [12]. A Canadian survey of parents of 5-17 year olds found children were less active and engaged in more screen time during the initial COVID-19 outbreak compared with before restrictions. Parent encouragement, participation and support of physical activity were found to be important correlates of children’s physical activity [21]. Zecevic et al [22] showed that, in circumstances not associated with lockdown, young children who receive greater parental support for physical activity were 6.3 times more likely to be highly active than inactive (B = 1.44, P<.05). In our study some parents made considerable efforts to keep their child physically active and thus minimise the impact of the restrictions. Many parents took advantage of the permission to leave the house for exercise to keep their child active. Yet despite these efforts, barriers specific to the lockdown were reported to reduce children’s physical activity, mainly relating to childcare and other facilities (such as playgrounds) being shut, as well as reduced contact with friends and family.

Our study identified disruptions to normal eating habits and routines, which varied by child and family. There were reports for some in changes in snacking, frequency of eating, volume of eating, eating out and who children were eating with. While some families speculated these changes would be temporary and would return to normal when usual childcare and socialisation resumed, it may be difficult to re-establish boundaries and break new habits [23,24].

One parent in our study discussed how free meals for her child at nursery had ceased. While our study did not identify acute food poverty, it is likely that pre-school children will have experienced food poverty in families where free school meals were no longer available and where the pandemic impacted family finances [25].

Rundle et al (2020) suggested the closure of schools during this period may exacerbate childhood obesity levels and increase disparities in risk of obesity for school-aged children [26]. While few parents mentioned concerns about weight gain, our findings suggest there is potential for obesity prevalence in pre-school children (and specifically children starting school in 2020) to have increased due to increased snacking and screen time, and decreased physical activity, particularly if some of the changes during lockdown have persisted. Annual height and weight data collection through the National Child Measurement Programme was stopped in March 2020. The national directive for it to resume in January 2021 is to be welcomed; this will enable assessment of the impact on children who have started school in September [27].

The negative impacts of COVID-19 for children living in poverty are expected to be the greatest [28,29]Our interviews sought to explore potential differences in the impact of lockdown across the socioeconomic spectrum. Half our participants lived in the most deprived IMD quintile, giving insight into the impact on families living in the areas of highest deprivation. We noted some differences within our sample. For example, the two families without access to gardens reflected on the difficult in keeping children active. In addition, families where parents were not working found it easier to engage children in physical activity than where parents were working. Beyond this, however, we did not note any obvious differences in narratives between those living in more or less deprived neighbourhoods. Overall, the impact of lockdown on children’s eating, physical activity and sedentary behaviours was felt similarly across the socioeconomic spectrum. The lack of difference could be due to characteristics of the sample; most families still had at least one parent in paid work and most had access to a garden. Interviews took place early in the COVID-19 crisis, so we may not yet be seeing the full effect, for example with the protective effect of the furlough scheme still in place.

This study raises the question of how parents can be supported during future lockdowns or local restrictions. The most recent periods of national lockdown in England and Wales, during the autumn of 2020, have not been as restrictive, with childcare, schools and some workplaces open. However, there are new and additional challenges: shorter, colder days, weariness of the restrictions, and increased financial pressures for some families with unemployment and redundancy rates at the highest seen for three and 12 years respectively [30]. In addition, positive cases in a childcare or school setting can result in an entire class (or ‘bubble’) being required to isolate for 14 days without being allowed out. The importance of supporting families to maintain or increase activity for young children within the home and in accessible outdoor environments becomes more prominent. Families need support to establish healthy revised routines and manageable healthy rules for snacks and screen time during periods of restriction. This needs to be presented in a context of supporting parents and not adding guilt or burden to parents during a period of stress. As highlighted in the recent Ipsos Mori report, which included an online survey of 1,000 parents of 0-5s in October 2020, 70% of parents reported feeling judged by others and almost half felt this negatively impacted on their mental health [31]. There is a role here for all sectors of health, social care and education including health visitors, children’s centres, early years providers, GPs, social services, local authority public health teams and Public Health England. Further, there is a role for research with pre-schoolers and 2020 school starters to be expanded [32] to understand the impact on their physical and social development, and particularly their risk of obesity, as they start school.

### Strengths and limitations

In-depth interviews enabled us to gain novel insights into the experiences of parents and pre-school children during lockdown and restrictions, although we acknowledge the limitation of investigating pre-school children’s experiences by asking their parents, rather than involving pre-school children directly. A strength of the study was in the ethnic and socioeconomic diversity of the sample from two regions of England, although this was a relatively educated sample (65% educated to degree level). In addition, we recognise that parents volunteering for interview may have had more interest in the topic or may have had different experiences to those that did not volunteer. Rigour was achieved by detailed data analysis and analytical decisions being shared with all team members to achieve credibility.

## Conclusions

While some positive changes were reported, there were wide-spread examples of lack of routines, habits and boundaries which, at least in the short-term, were likely to have been detrimental for child health and development. Guidance and support for parents and families on how to maintain healthy routines and compensate for future COVID-19 restrictions could be valuable to protect child health and ensure that unhealthy habits are not adopted.

## Data Availability

Anonymised study data will be made available via a University of Bristol repository

## Abbreviations

BAME: Black, Asian and Minority Ethnic
CO-SPYCE: Covid-19: Supporting Parents and Young Children during Epidemics (study)
COVID-19: Coronavirus Disease (also known as SARS-CoV-2)
GPs: General Practitioners
IMD: Index of Multiple Deprivation
UK: United Kingdom

## Ethics approval and consent to participate

Ethical approval for the study was granted by University of Bristol Faculty of Health Sciences Research Ethics Committee. Participant consent was recorded on a separate audio file before the interview commenced.

### Consent for publication

Consent to include anonymised information in publications was given by all participants.

### Availability of data and materials

Anonymised study data will be made available via a University of Bristol repository

## Authors’ contributions

The study was conceived by JC, RK, SC, KW, HT, RB, KH, SS and RL. JC led the study with oversight from RL and RK. Interviews were conducted by JC, SC and KW. Coding of the data was performed by JC, HT and RB. JC, RK and RL produced the first draft of the manuscript, with all other authors providing critical review and intellectual content. All authors read and approved the final manuscript.

## Acknowledgements

We are very grateful to all the parents who took part in the research interviews. The views expressed in this publication are those of the authors and not necessarily any of the funding bodies listed.

